# On short-term trends and predictions for COVID-19 in France and the USA: comparison with Australia

**DOI:** 10.1101/2020.11.17.20233718

**Authors:** Henry C. Tuckwell, Mohsen Dorraki, Stephen J. Salamon, Andrew Allison, Derek Abbott

## Abstract

In Europe and the USA daily new COVID-19 cases have recently been occurring in record numbers, which has created an alarming situation. The CDC in conjunction with several University groups gives forecasts for each county in the USA for several weeks at a time, but they have very large confidence intervals typified by the most recent national prediction of between 310,000 and 710,000 new cases for the week ending November 21, 2020. We have examined recent data for France and the USA over 10, 15 and 20 days. Using such data with simple fitting techniques, which do not require knowledge of any parameters, it has been possible to predict new case numbers fairly accurately for a week or more. A best-fitting polynomial of high order was only useful for a few days, after which it severely overestimated case numbers. A more detailed analysis with confidence intervals was performed for polynomials of orders one to six, which showed that lower order polynomials were more useful for prediction. Using the packages PCHIP and a POLYFIT (with degree one) in MATLAB gave smooth curves from which future case numbers could be reasonably well estimated. With PCHIP the average errors over 7 days were remarkably small, being −0.16% for France and +0.19% for the USA. A comparison is made between the temporal patterns of new cases for France, the USA and Australia. For Australia the second wave has dwindled to close to zero due to hard lock down conditions, which are discussed.

## 1 Introduction

There have been numerous articles on modeling and predicting COVID-19 case numbers since the beginning of the pandemic, which started in Wuhan, China in December 2019 (Nunes et al., 2020; Roda et al., 2020). This disease is readily transmitted by the SARS-CoV-2 virus. Some important characteristics of infection are (i) the incubation period, which is the time from infection to the appearance of symptoms, (ii) the latent period, which is the time from infection to becoming infectious or contagious, and (iii) the infectious period. Typical durations are five to six days for the incubation period, a further two days to enter the infectious period which lasts approximately 10 days (Lauer et al., 2020; Roda et al., 2020). However, the infectious period may commence before the end of the incubation period (Liu et al., 2020).

Data are available at several internet sites for the daily numbers of new cases and various other summary statistics for each country. The definition of new case however is rather imprecise as it could be from observations of appearance of symptoms or the result of laboratory testing.

Modeling in the context of COVID-19 has been of various types. Wynants et al. (2020) reviewed the literature on the medical aspects of the disease to May 5, 2020 and found four studies which identified subjects at risk, 91 on the detection of the virus and 50 prognostic models. However, the term epidemic modeling usually refers to mathematical models which are used to attempt to predict the time course of the numbers of infected individuals in a population. Possibly the first such modeling, concerning life expectancy after smallpox infection, seems to have been that of Bernoulli (1766) as described by Dietz and Heesterbeek (2002). Another pioneering contribution was the SIR (Susceptible, Infected, Recovered) differential equation model of Kermack and McKendrick (1927).

The CDC (Centers for Disease Control and Prevention) in the USA co-ordinates the results of 32 modeling groups based mainly in US Universities.They list graphical and numerical forecasts by county for USA^1^. In their most recent 4-week forecast for the whole country it was found likely that there would be between 310,000 and 710,000 new cases in the week ending November 21, 2020. They cautioned that there were uncertainties in the predictions and that recent observations had fallen outside the confidence bands, correlating with new record daily numbers of cases. Chen et al. (2020) consider a model which takes into account 51 compartments, representing US states plus District of Columbia. Within each compartment there are five groups of individuals called susceptible (S), exposed (E), reported (I), unreported (A) and resolved (R) with numbers evolving according to ordinary differential equations. These authors found that interstate travel in the USA was not a major factor in the spread of infections.

Bardina et al. (2020) use an SIR model containing 12 parameters with latent and quarantine periods which extends the simpler model of Tuckwell and Williams (2007) and that of Ferrante et al. (2016). They investigate the effects of the length of the quarantine period and the infectivity of asymptomatic individuals on the time course of the epidemic. Linka et al. (2020) develop an SEIR model with time-varying reproductive number to analyse the growth of COVID-19 numbers across Europe.

Other authors, including Wu et al. (2020), Roda et al. (2020) and Roosa et al. (2020) focus on the spread of the disease from its first epicenter in the Chinese province of Wuhan. Wu et al. (2020) analyse early numbers of cases exported internationally to estimate the number of infected in Wuhan and subsequently the numbers of cases exported from Wuhan to other Chinese cities. Roda et al.(2020) also focus on the early data and employed both SIR and SEIR models to forecast the number of cases. The simpler SIR model performs better and also has the advantage of fewer parameters. Roosa et al. (2020) assume several underlying growth models, including a generalized logistic model, to predict the number of COVID-19 cases in China for 5, 10 and 15 days in February 2020.

Zhang et al. (2020) develop a detailed stochastic model for the COVID-19 epidemic in China. Chang et al. (2020) study the COVID-19 outbreak in Australia using a simulator called AceMod, which had been previously used for influenza and incorporates the census demographic data for the country’s whole population. Nunes et al. (2020) review an array of approaches to modeling the COVID-19 pandemic with a discussion of their merits, while Gomes (2020) focuses on its development in Portugal where initial estimates were that 85% of the population would be infected before the epidemic ended. Colombo et al. (2020), modifying the approach of Flaxman et al. (2020), ascertain the effect of a heterogeneous versus homogeneous population of susceptibles on modeling for the United Kingdom. Zeng and Ghanem (2020) employ the method of switching Kalman filters to predict COVID-19 cases in the USA. Their article also contains a summary of, and references to, several previously used techniques. Deep learning techniques are also used not only for virological analysis (Zhu et al., 2020) but also to predict COVID-19 case numbers in India (Arora et al., 2020).

## 2 Methods and Results

Data on the numbers of daily new cases for France, the USA and Australia were obtained for the period January 1 to November 3, 2020 from the downloadable daily postings at the European Centre for Disease Prevention and Control’s website^2^.

The daily increments were converted to cumulative data and parts of the resultant curves over the 20 day period to October 28, 2020 are shown in Figure 1. The maxima for France and the USA are respectively 1,198,695 and 8,779,653 which occur on day 20 of these sets, whereas the maximum for Australia is 31 on day 15. If these maxima are scaled by population size, the value for France is about 14,277 times that of Australia whereas for the USA the maximum is about 7.3 times that of France and 104,570 times that of Australia. These figures underscore just how alarming the numbers of cases are in France and the USA. The plots for France, the USA and Australia have distinctly different forms - see Section 3.1 for an overview of the overall time-courses. It is clear that mathematical models for France and the USA would need to be very different from a model to fit the Australian data.

**Figure 1:**
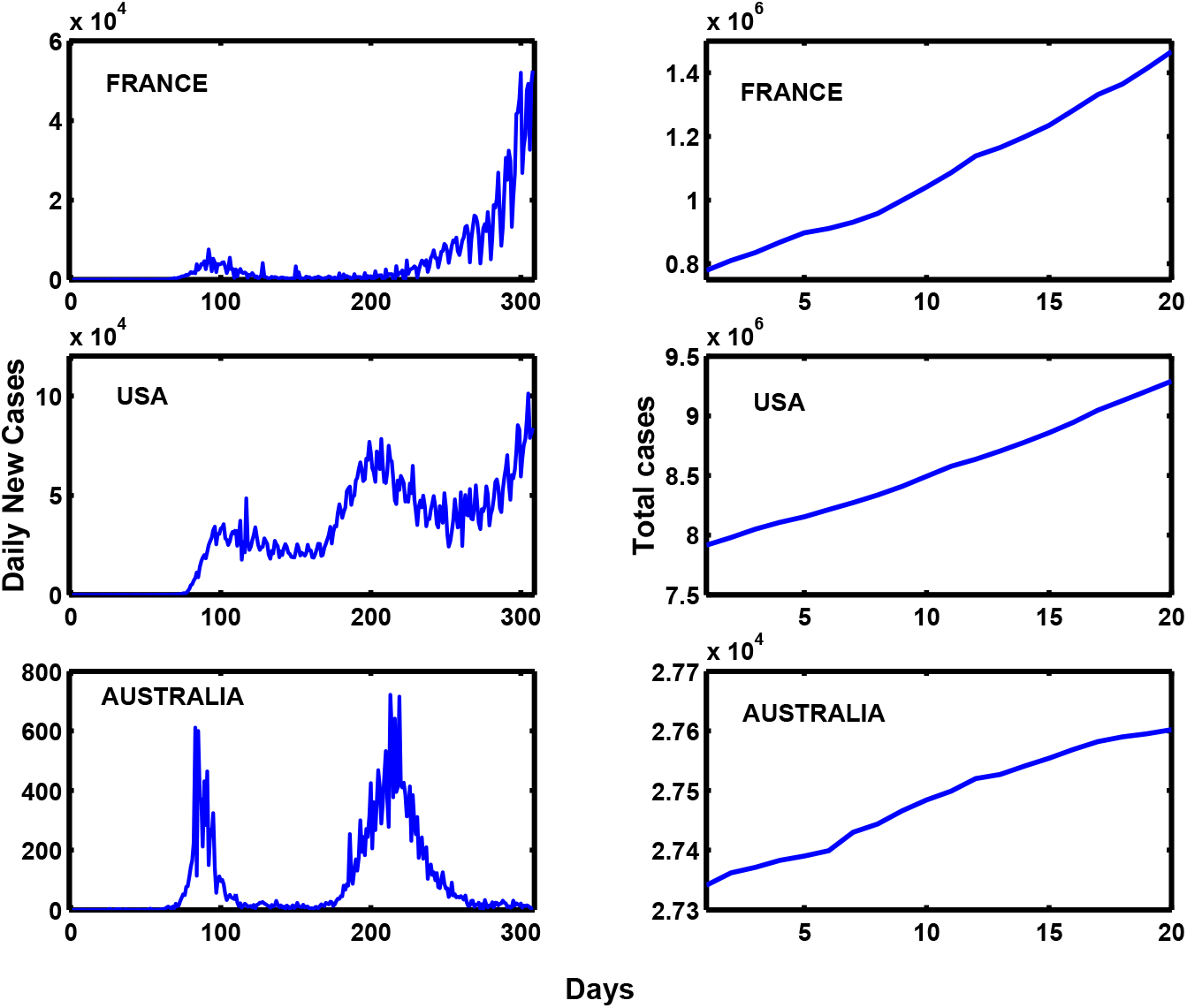
Left column. Daily new COVID-19 cases in France, USA and Australia for January 1 - November 3, 2020. Right column. Total COVID-19 cases in France, USA and Australia for 20 days ending on November 3, 2020.

### 2.1 Fitting with polynomials

One method of trying to fit data such as those for COVID-19 for France and the USA is to obtain a best-fitting polynomial from the given data and then use the polynomial to etrapolate to future times. We first carry out this multiple linear regression procedure using MATLAB’s basic fitting program POLYFIT. The polynomial fitting the French data with the lowest norm of residuals is the following of 6th degree

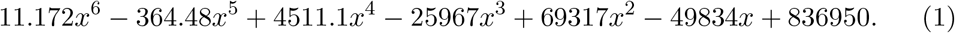

This function is plotted over the data interval of 10 days along with the data in Figure 2A and is seen to fit the data well. However, when plotted over the extended interval of 17 days as in Figure 2B, for the first few days past the data interval the disagreement between the polynomial and a plausible continuation of the data is mild, but then rapidly escalates to values that are clearly unrealistic. This points to the conclusion that this method of choosing a polynomial for extrapolation of the COVID-19 data is not accurate.

**Figure 2:**
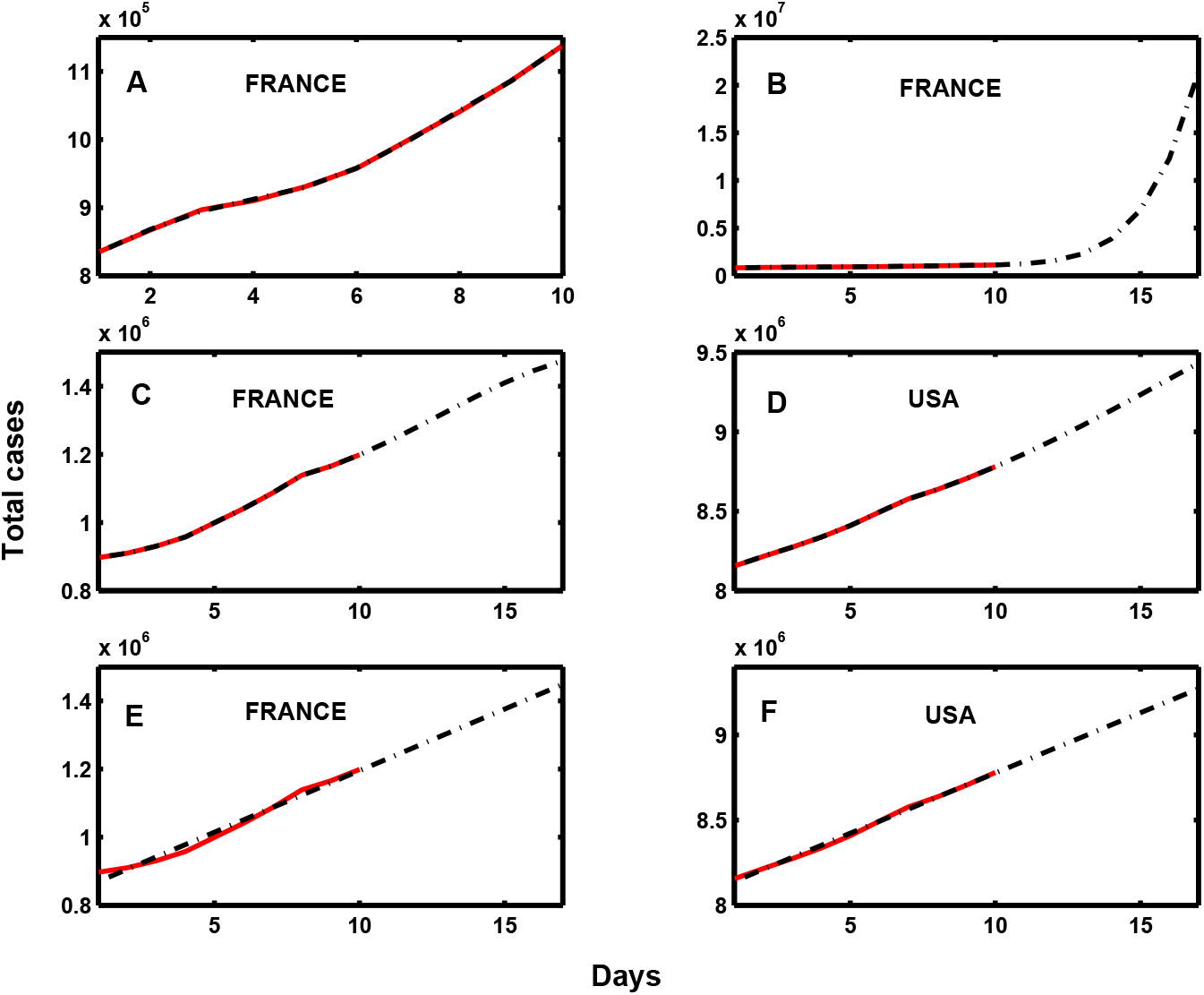
(A). Data for France for 10 days from October 17 to October 26, 2020 (red) and best-fitting sixth-degree polynomial (black dash-dot). (B). Case numbers for France from October 17 to November 2, 2020 (black dash-dot) using extrapolation with the best-fitting sixth-degree polynomial, superimposed on the data in A. (C). Data for France for 10 days from October 19 to October 28, 2020 (red). PCHIP was used to fit the data and extrapolate to November 4, 2020 (black dash-dot). (D). As in C but for USA. (E) and (F). As in C and D using least squares regression straight line to fit and extrapolate the data.

However, repeating this ordinary least squares (OLS) regression fitting of polynomials with our own software, this 6th order polynomial appears to be a case of overfitting, from the point-of-view of prediction accuracy. This may be demonstrated by cross-validation, calculating the PRESS statistic, the sum of squared differences between each observation and a polynomial fitted to all the other observations. The 6th order polynomial has 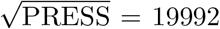, while arguably the best fit is the 4th order polynomial with 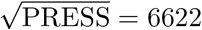. High order polynomial fits are problematic for extrapolation beyond the region of the observed data, as uncertainty in the high order coefficients leads to very large confidence intervals. Hence a low order polynomial (1st or 2nd order) may be preferred for predictions beyond the known data. This is demonstrated in the plots for 1st to 6th order polynomials in Figure 3.

**Figure 3:**
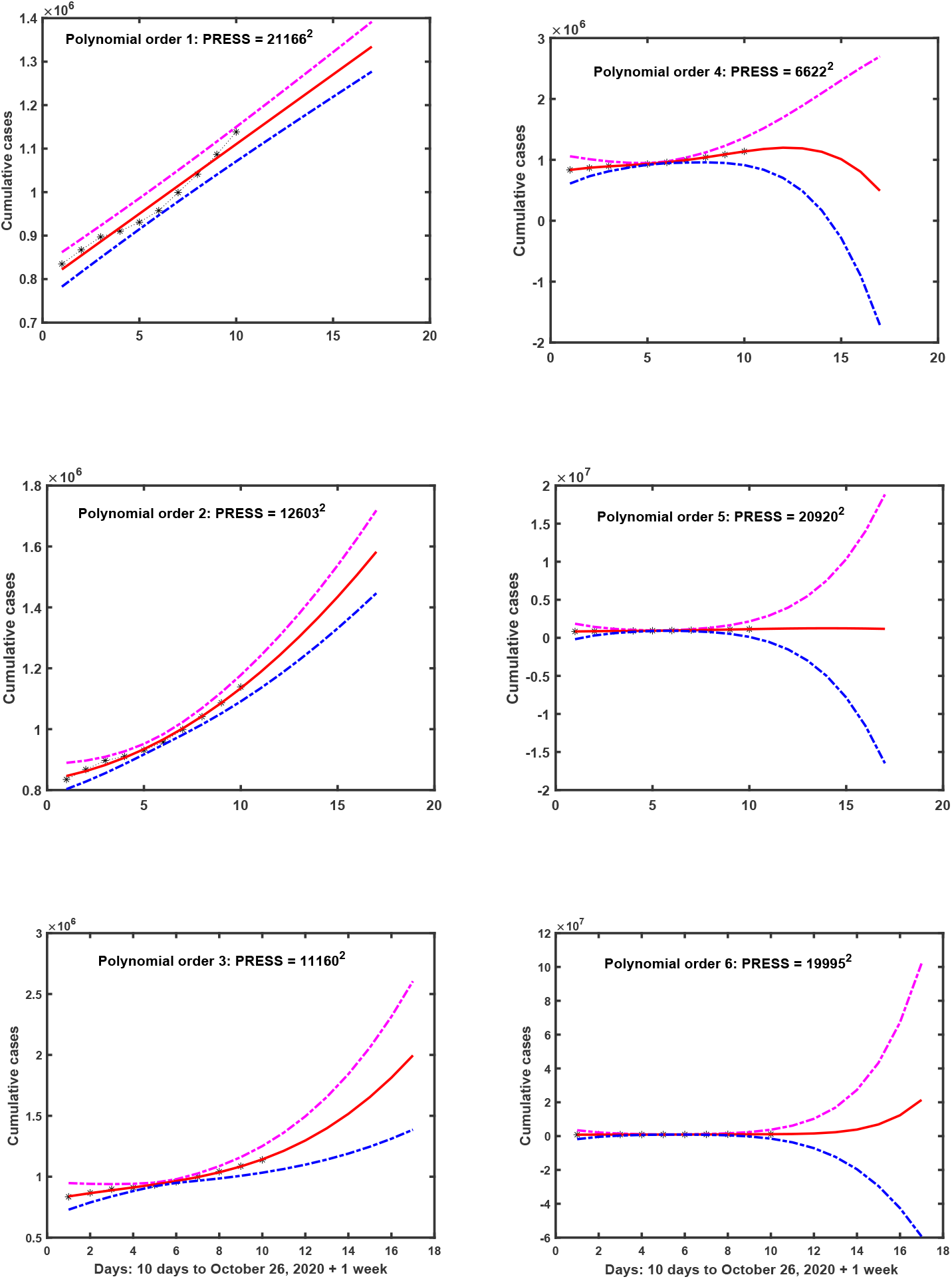
Plots showing the data for France for 10 days (black) and best fitting polynomials (red), including extrapolation for another week. The polynomials are from order 1 to 6 and 95% confidence interval limits are shown (magenta and blue dash-dot lines). The confidence intervals are extreme for high orders due to estimated standard error in the high order coefficients of the polynomials.

### 2.2 Using PCHIP

The MATLAB program called PCHIP (Piecewise Cubic Hermite Interpolating Polynomial) is primarily a tool for interpolation at points between data points but as suggested by the documentation, it can also be used to extrapolate as shown in a successful extrapolation of US census data^3^.

This method was used to extend the COVID-19 data for France and the USA using previous data for 10, 15 and 20 days to obtain estimates for a week ahead. The results on using data for 10 days are shown in Figures 2C and 2D, the fitted curve being shown for the whole 17 days. We find that the results are the same whether data for 10, 15 or 20 previous days is employed. The numerical results for the seven extra days are shown in Tables 1 and 2, second column.

**Table 1:** Predicted cases for France for the week after October 28, 2020 from previous 10 days data. Column five gives the data for Oct 29-Nov 4. Error % is for PCHIP relative to Data. Simple LR = simple linear regression.

| Day, Date | PCHIP | % Error | Simple LR | Data |
| --- | --- | --- | --- | --- |
| 1, Oct 29 | 1,238,000 | +0.23 | 1,231,400 | 1,235,132 |
| 2, Oct 30 | 1,281,100 | -0.12 | 1,267,600 | 1,282,769 |
| 3, Oct 31 | 1,325,600 | +0.48 | 1,303,700 | 1,331,984 |
| 4, Nov 1 | 1,369,500 | +0.36 | 1,339,900 | 1,364,625 |
| 5, Nov 2 | 1,410,400 | -0.24 | 1,376,100 | 1,413,915 |
| 6, Nov 3 | 1,446,300 | -0.01 | 1,412,200 | 1,466,433 |
| 7, Nov 4 | 1,474,900 | -1.85 | 1,448,400 | 1,502,763 |

**Table 2:** Predicted cases for the USA for the week after October 28, 2020 from previous 10 days data. Column five gives the data for Oct 29-Nov 4. Error % is for PCHIP relative to Data. Simple LR = simple linear regression.

| Day | PCHIP | % Error | Simple LR | Data |
| --- | --- | --- | --- | --- |
| 1, Oct 29 | 8,861,200 | +0.03 | 8,847,000 | 8,858,024 |
| 2, Oct 30 | 8,948,300 | +0.02 | 8,917,700 | 8,946,154 |
| 3, Oct 31 | 9,039,900 | -0.08 | 8,988,500 | 9,047,427 |
| 4, Nov 1 | 9,135,100 | +0.1 | 9,059,200 | 9,126,361 |
| 5, Nov 2 | 9,232,900 | +0.28 | 9,130,000 | 9,207,362 |
| 6, Nov 3 | 9,332,400 | +0.44 | 9,200,700 | 9,291,245 |
| 7, Nov 4 | 9,432,500 | +0.51 | 9,271,500 | 9,383,979 |

### 2.3 Using simple linear regression

Noting that the data and its PCHIP extrapolation seem to lie practically on a straight line, we employ the POLYFIT program to find a best fitting polynomial of degree one, which is equivalent to a least squares straight line or regression stgraight line. The results on extrapolating the data to 7 days ahead are shown for France and the USA in Figures 2E and 2F. The numerical results are given in Tables 1 and 2, 4th column.

## 3 Discussion

There are many deterministic and stochastic models for predicting the time courses of epidemics, but it is difficult to apply them to real data because of the complexities of both the host biological populations and the dynamics of agents of disease such as bacteria or viruses. Another complication in studying the spread of disease in human populations is that the hosts may change their behavior, voluntarily or mandated, so that many of the parameters of a model are variable. Such changes with respect to COVID-19 include curfews, closure of schools and businesses, quarantine, social distancing, face masks, travel restrictions, hand washing, and vaccination when available. Furthermore, the number of new cases reported daily depends, in an uncertain way on the number of tests.

The wide disparities in the time courses of COVID-19 infection in various countries (Nunes et al., 2020) supports the view that it is likely to be impracticable to develop accurate models of either a deterministic or stochastic ilk. This diversity of time courses is highlighted in Figure 1, where the daily new cases for France, the USA and Australia are plotted. Broadly speaking, France and Australia show evidence of first and second waves, whereas the USA exhibits three waves. For Australia the second wave seems to have subsided to almost negligible levels, whereas in France the number of daily cases in the second wave is still increasing.

### 3.1 The time course for Australia

In May and the first half of June, 2020, the daily numbers of new cases of COVID-19 in Australia were very small so that it looked like there would be no second wave. For most of Australia there was in fact no significant second wave wave, but in Melbourne in the state of Victoria some breaches in quarantine protocol were reported. Consequently a second wave grew very rapidly from late June to early August with daily new cases rising to over 700. Severe lock down measures finally brought the numbers down to pre-second wave levels by the end of September.

The conditions of this 4th stage lock down were

- Compulsory mask wearing outside the home
- Curfews from 9 pm to 5 am every day
- Travel more than 5 km from home forbidden
- All shops but not supermarkets closed
- Schools, tertiary education institutions and businesses including restaurants closed
- Supermarket shopping by one person per household only
- Social distancing unbiquitous with separation 1.5 to 2 m
- Exercise for no more than an hour, near home, is allowed
- Maximum 5 people at a wedding and 10 at a funeral. Limited numbers at social gatherings.
- Large fines imposed if rules broken

The severe lockdown conditons applied only to Melbourne and its suburbs, not regional Victoria which had seen relatively very few cases during the second wave. Nor did they apply to any of the other seven states or territories of Australia which had negligible numbers of COVID-19 cases. It is apparent that even though the second wave should have been avoided, the 4th stage lockdown did a good job at eventually driving the number of cases down. It is possible that if such lockdown conditions, including the threat and enforcement of financial penalties, were applied in Europe and the USA, the numbers of COVID-19 cases there would drop substantially.

### 3.2 Concluding remarks

We have demonstrated that short term predictions for about a week and possibly longer may be successfully carried out with readily available software programs. The program PCHIP in MATLAB gave remarkably accurate predictions for France and the USA for at least a week. However, during the time period examined, the time courses of the epidemics vary smoothly. If significant changes in rate of change occur, which can be detected using standard trend tests, then the data must be broken up into appropriate segments.

## Data Availability

All data is available on the internet as given in the links below.

https://www.cdc.gov/coronavirus/2019-ncov/cases-updates/forecasts-cases.html

https://www.ecdc.europa.eu/en/publications-data/download-todays-data-geographic-distribution-covid-19-cases-worldwide

## Footnotes

1 https://www.cdc.gov/coronavirus/2019-ncov/cases-updates/forecasts-cases.html

2 https://www.ecdc.europa.eu/en/publications-data/download-todays-data-geographic-distribution-covid-19-cases-worldwide

3 https://www.mathworks.com/company/newsletters/articles/fitting-and-extrapolating-u-s-census-data.html

